# Evidence and magnitude of seasonality in SARS-CoV-2 transmission: Penny wise, pandemic foolish?

**DOI:** 10.1101/2020.08.17.20176610

**Authors:** Adam Kaplin, Caesar Junker, Anupama Kumar, Mary Anne de Amorim Ribeiro, Eileen Yu, Michael Wang, Ted Smith, Shesh N Rai, Aruni Bhatnagar

## Abstract

**Importance:** Intensity and duration of the COVID-19 pandemic, and planning required to balance concerns of saving lives and avoiding economic collapse, could depend significantly on whether SARS-CoV-2 transmission is sensitive to seasonal changes.

**Objective:** Hypothesis is that increasing temperature results in reduced SARS CoV-2 transmission and may help slow the increase of cases over time.

**Setting:** Fifty representative Northern Hemisphere countries meeting specific criteria had sufficient COVID-19 case and meteorological data for analysis.

**Methods:** Regression was used to find relationship between the log of number of COVID-19 cases and temperature over time in 50 representative countries. To summarize the day-day variability, and reduce dimensionality, we selected a robust measure, Coefficient of Time (CT), for each location. The resulting regression coefficients were then used in a multivariable regression against meteorological, country-level and demographic covariates.

**Results:** Median minimum daily temperature showed the strongest correlation with the reciprocal of CT (which can be considered as a rate associated with doubling time) for confirmed cases (adjusted R^2^ = 0.610, p = 1.45E-06). A similar correlation was found using median daily dewpoint, which was highly colinear with temperature, and therefore was not used in the analysis. The correlation between minimum median temperature and the rate of increase of the log of confirmed cases was 47% and 45% greater than for cases of death and recovered cases of COVID-19, respectively. This suggests the primary influence of temperature on SARS-CoV2 transmission more than COVID-19 morbidity. Based on the correlation between temperature and the rate of increase in COVID-19, it can be estimated that, between the range of 30 to 100 degrees Fahrenheit, a one degree increase leads to a 1% decrease--and a one degree decrease leads to a 3.7% increase--in the rate of increase of the log of daily confirmed cases.

**Conclusion:** The results suggest that boreal summer months are associated with slower rates of COVID-19 transmission, with the reverse true in winter months. Knowledge of COVID-19 seasonality could prove useful in local planning for phased reductions social interventions and help to prepare for the timing of possible pandemic resurgence during cooler months.

## Introduction

Severe Acute Respiratory Syndrome Coronavirus 2 (SARS-CoV-2), is a newly-identified enveloped, non-segmented, positive sense RNA virus. It is responsible for Corona VIrus Disease 2019 (COVID-2019), which was designated a pandemic by the World Health Organization (WHO) on March 11, 2020.(1) SARS-CoV-2 belongs to a large family of human corona viruses such as MER-CoV and SARS-CoV, which are respiratory pathogens associated with a range of respiratory and non-respiratory outcomes. Like other respiratory infectious agents, coronaviruses display marked seasonality.(2) Infection rates peak in mid-winter, and then diminish during summer months. Because SARSCoV-2 is a recently identified human pathogen, seasonal variations in its transmission have not become evident, though there is much speculation that seasonal change with the boreal summer might decrease infection rates of SARS-CoV-2 and help flatten the epidemic curve of COVID-19.(3)

A better understanding of the seasonal dependence of SARS-CoV-2 infections, and the identification of environmental conditions that regulate its spread, would be helpful in assessing the differential impact of the virus in different geographic locations under various meteorological conditions. Such knowledge would also help in predicting the timing and degree of potential slowing of transmission in the summer and resurgence of the virus during winter.(4)

To understand the seasonal dependence of SARSCoV-2 infections in the absence of year-long incidence data, we examined the effect of atmospheric temperature, the main variable with season,(4) on SARS CoV-2 transmissibility using currently available data on the rates of SARS-CoV-2 infection during winter and spring from fifty representative countries in the Northern Hemisphere.

## Methods

Longitudinal changes in cumulative COVID-19 confirmed deaths, and recovered cases from 1/22/20 through 4/6/20 were estimated from databases made available by the Johns Hopkins University.(5) The start date of January 22, 2020 was chosen because that is when the database started tracking cases. For analysis, we selected only the Northern Hemisphere countries because even though the number of cases have been increasing on both sides of the equator, only the Northern Hemisphere experienced an increase in temperature during the study period. The opposite was true in the Southern Hemisphere. As a result, the impact of temperature on the rate of new cases cannot be investigated simultaneously on pooled data from both hemispheres. We excluded from analysis China(6) and Russia,(7) and countries where Russian was the primary language, because of inconsistent reporting over time. Italy was excluded because number of confirmed cases seemed significantly larger than adjacent countries for reasons that are not entirely clear.(8)

To preserve methodological conformity, countries with separate reports of cases from multiple regions were excluded. Other countries were excluded due to limited access to, or the lack of, reliable data on COVID-19 cases or meteorological data. To enable analysis of cases during consecutive days in a log linear fashion, we established the following criteria to ensure sufficient numbers of confirmed cases: to be included during time period 1/22/20-4/6/20, countries needed to have a median number of confirmed cases > 1 or the mean number of confirmed cases > 75.

The resulting 50 representative countries (Table 1) had a mean latitude of 38 +/−15 SD, range 1–65 degrees, that covers 70% of the total range (1-90 degrees), with each country being separated from the next by a mean of 1.3 +/−1.7 SD degrees. Countries mean longitude as 34+/−44 SD, range −103 to 138 that covered 70% of the total range (−180 to 180 degrees), with each country separated from the next by a mean of 6 +/− 13 SD degrees. The resulting wide range of temperatures across the 50 countries was as follows: 1) mean average temperature 51.1 +/−16.9 °F, median 44.9 °F, range 22.6 – 85.7 °F, and 2) mean minimal temperature 42.0+/−16.3 °F, median 36.6 °F, range 15.5 – 77 °F.

**Table 1:** First Regression: Rate of rise of confirmed cases of COVID-19 in 50 countries, and the associated local meteorological and demographic variables.

| Country | CT Conf Cases | CT<sup>-1</sup> Conf Cases | R<sup>2</sup> | Tmin Median | DEWP Median | Land Area Per Capita | Median age | Conf Cases Days |
| --- | --- | --- | --- | --- | --- | --- | --- | --- |
| Algeria | 0.081 | 12.42 | 0.969 | 56.3 | 25.2 | 0.05431 | 28.9 | 42.0 |
| Armenia | 0.103 | 9.74 | 0.909 | 26.6 | 25.2 | 0.00961 | 36.6 | 37.0 |
| Austria | 0.097 | 10.26 | 0.941 | 29.1 | 31.2 | 0.00915 | 44.5 | 42.0 |
| Belgium | 0.090 | 11.17 | 0.913 | 38.7 | 37.4 | 0.00261 | 41.6 | 63.0 |
| Bosnia and Herzegovina | 0.086 | 11.68 | 0.967 | 20.3 | 23.8 | 0.01554 | 43.3 | 33.0 |
| Croatia | 0.074 | 13.59 | 0.977 | 32.9 | 32.8 | 0.01363 | 43.9 | 42.0 |
| Czechia | 0.091 | 10.93 | 0.942 | 33.0 | 30.2 | 0.00721 | 43.3 | 37.0 |
| Egypt | 0.076 | 13.21 | 0.928 | 48.3 | 38.7 | 0.00973 | 24.1 | 53.0 |
| Estonia | 0.088 | 11.36 | 0.908 | 31.5 | 32.5 | 0.03196 | 43.7 | 40.0 |
| Finland | 0.062 | 16.13 | 0.899 | 19.3 | 22.3 | 0.05485 | 42.8 | 69.0 |
| France | 0.071 | 14.03 | 0.940 | 34.6 | 38.2 | 0.00839 | 41.7 | 74.0 |
| Germany | 0.074 | 13.49 | 0.940 | 34.7 | 33.4 | 0.00416 | 47.8 | 71.0 |
| Greece | 0.074 | 13.52 | 0.904 | 40.1 | 42.2 | 0.01237 | 45.3 | 41.0 |
| Hungary | 0.081 | 12.29 | 0.965 | 32.0 | 31.3 | 0.00937 | 43.6 | 34.0 |
| Iceland | 0.072 | 13.81 | 0.881 | 15.5 | 17.4 | 0.29378 | 37.1 | 39.0 |
| India | 0.055 | 18.32 | 0.903 | 64.0 | 56.6 | 0.00215 | 28.7 | 68.0 |
| Iran | 0.076 | 13.18 | 0.820 | 34.8 | 18.5 | 0.01939 | 31.7 | 48.0 |
| Iraq | 0.058 | 17.13 | 0.913 | 49.3 | 45.5 | 0.01080 | 21.2 | 43.0 |
| Ireland | 0.104 | 9.66 | 0.947 | 35.8 | 37.4 | 0.01395 | 37.8 | 38.0 |
| Israel | 0.096 | 10.45 | 0.986 | 46.0 | 42.3 | 0.00250 | 30.4 | 46.0 |
| Japan | 0.041 | 24.67 | 0.967 | 44.6 | 38.2 | 0.00288 | 48.6 | 76.0 |
| Kuwait | 0.035 | 28.56 | 0.792 | 56.9 | 46.9 | 0.00417 | 43.2 | 43.0 |
| Lebanon | 0.067 | 15.02 | 0.929 | 41.8 | 40.4 | 0.00150 | 29.7 | 46.0 |
| Malaysia | 0.044 | 22.77 | 0.932 | 74.8 | 75.8 | 0.01015 | 33.7 | 73.0 |
| Mexico | 0.089 | 11.23 | 0.967 | 46.4 | 36.7 | 0.01508 | 29.2 | 39.0 |
| Moldova | 0.098 | 10.23 | 0.947 | 31.6 | 29.3 | 0.00814 | 29.3 | 30.0 |
| Morocco | 0.098 | 10.19 | 0.974 | 44.2 | 18.5 | 0.01209 | 37.7 | 36.0 |
| Netherlands | 0.097 | 10.34 | 0.894 | 36.8 | 37.5 | 0.00197 | 29.1 | 40.0 |
| Norway | 0.080 | 12.50 | 0.840 | 21.1 | 21.5 | 0.06738 | 42.8 | 41.0 |
| Oman | 0.049 | 20.28 | 0.972 | 57.9 | 52.9 | 0.06061 | 39.5 | 43.0 |
| Pakistan | 0.095 | 10.51 | 0.952 | 49.6 | 36.5 | 0.00349 | 26.2 | 41.0 |
| Philippines | 0.055 | 18.09 | 0.863 | 74.2 | 76.1 | 0.00272 | 30.1 | 68.0 |
| Poland | 0.102 | 9.80 | 0.923 | 31.1 | 30.9 | 0.00809 | 24.1 | 34.0 |
| Portugal | 0.109 | 9.16 | 0.957 | 47.9 | 48.6 | 0.00898 | 41.9 | 36.0 |
| Qatar | 0.056 | 17.82 | 0.798 | 57.9 | 49.5 | 0.00403 | 33.7 | 38.0 |
| Romania | 0.095 | 10.50 | 0.977 | 28.4 | 30.3 | 0.01196 | 44.6 | 41.0 |
| Saudi Arabia | 0.098 | 10.22 | 0.927 | 59.0 | 29.0 | 0.06175 | 42.5 | 36.0 |
| Serbia | 0.105 | 9.53 | 0.916 | 34.2 | 32.9 | 0.01001 | 30.8 | 32.0 |
| Singapore | 0.031 | 32.04 | 0.932 | 77.4 | 74.7 | 0.00012 | 43.4 | 75.0 |
| Slovakia | 0.079 | 12.65 | 0.841 | 26.5 | 27.4 | 0.00881 | 35.6 | 32.0 |
| Slovenia | 0.067 | 14.83 | 0.803 | 32.3 | 33.3 | 0.00969 | 44.9 | 33.0 |
| South Korea | 0.058 | 17.10 | 0.882 | 29.4 | 23.6 | 0.00190 | 44.9 | 76.0 |
| Spain | 0.100 | 10.01 | 0.949 | 42.2 | 43.0 | 0.01067 | 43.9 | 66.0 |
| Sweden | 0.076 | 13.14 | 0.915 | 18.9 | 22.8 | 0.04063 | 41.1 | 67.0 |
| Switzerland | 0.096 | 10.43 | 0.899 | 30.7 | 33.6 | 0.00457 | 42.7 | 42.0 |
| Thailand | 0.032 | 31.33 | 0.869 | 73.4 | 71.3 | 0.00732 | 39.0 | 76.0 |
| Turkey | 0.177 | 5.65 | 0.924 | 27.4 | 31.1 | 0.00913 | 32.2 | 27.0 |
| United Arab Emirates | 0.039 | 25.87 | 0.950 | 64.8 | 56.9 | 0.00845 | 38.4 | 69.0 |
| United Kingdom | 0.075 | 13.33 | 0.964 | 36.1 | 36.3 | 0.00356 | 40.6 | 67.0 |
| Vietnam | 0.027 | 36.94 | 0.924 | 73.2 | 71.4 | 0.00319 | 31.9 | 75.0 |

Daily weather data for each of the 50 countries were obtained from National Oceanic and Atmospheric Administration (NOAA).(9) Daily minimum temperatures (Tmin) were used because they appear to be more important for virus transmissibility.(10) We used Dew Point (DP) as a measure of humidity because it is an absolute assessment of water vapor content that does not depend upon temperature, and it is the only country meteorological data related to humidity that is tracked by NOAA. The daily Tmin and DP for each country were obtained from NOAA station locations closest to the specific latitude and longitude that the JHU database listed for each country. The medians of the daily Tmin and daily DP were derived for each country for whatever portion of 1/22/20 through 4/6/20 that they had cases of confirmed COVID-19 reported (Table 1). Median was chosen because it is less affected by outliers and skewed data than the mean.

## Results

### Analysis of the impact of changing temperature on the rate of rise of COVID-19 cases

To investigate the relationship between Tmin and COVID-19 transmissibility, we devised a two-step process. In the first step, starting with the date of the first case for each country we examined linear regressions of the log of the daily cumulative COVID-19 cases versus time in sequential days (Figure 1). From this analysis we calculated the resulting regression coefficients (which we termed coefficients of time (CT) with units of log cases/day) for each country (Table 1). To determine consecutive days, we converted calendar dates to sequential serial numbers that can be used in calculations—e.g. 1/22/2020 was converted to 43852 because it is 43,851 days after 1/1/1900. The CT of log confirmed cases vs sequential days for all fifty countries yielded a mean overall R^2^ of 0.919 +/− 0.049 SD, range 0.792–0.986, and p-value mean 4.20 ×10^−14^ ± 2.61 ×10^−13^, range 1.00 ×10^−15^ – 1.84 × 10^−12^ (Table 1).

**Figure 1:**
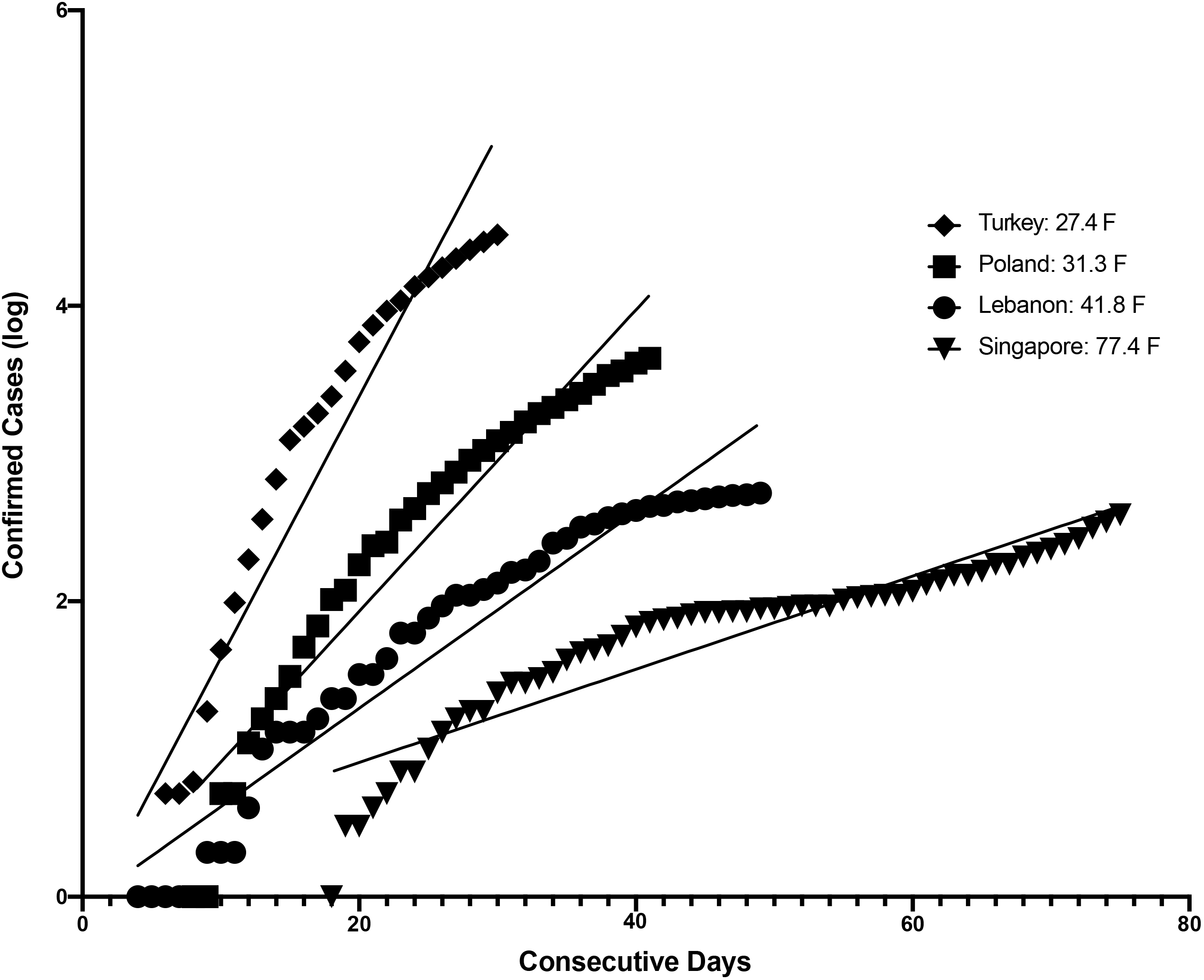
The first regression of log of confirmed cases vs consecutive days for 4 of the total of 50 countries is Shown. Starting with the date of the first case for each country, which was standardized to day 1 for purposes of visual comparison, we examined linear regressions of the log of the daily cumulative COVID-19 cases versus time in sequential days. From this analysis we calculated the resulting regression coefficients (which we termed coefficients of time (CT) with units of log cases/day) for each country.

In the second step, we investigated whether CT was associated with the median daily T_min_ (Tmin_Med) in each country for the same time period that cases had been reported. Because of the approximate proportionality of fitted CT with the standardized residuals when performing linear regression with Tmin_Med, we used a reciprocal transformation of the dependent variable. (11) Therefore, for the second regression for all countries we used the reciprocal of CT (CT^−1^) (Figure 2).

**Figure 2:**
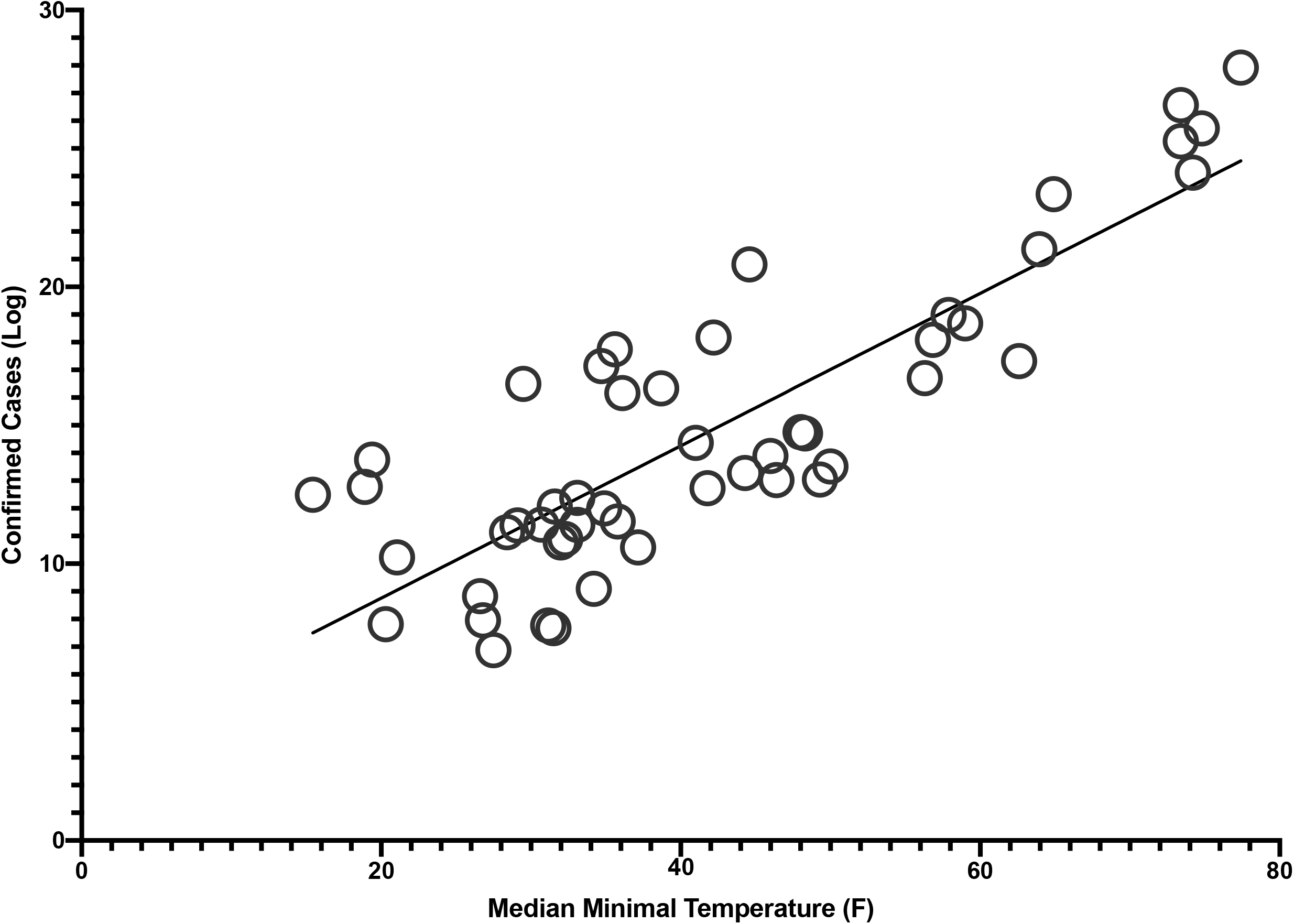
Shows for fifty representative countries the regression of the reciprocal of the rate of increase of the log of confirmed cases (CT^−1^) vs the median minimal temperature. This establishes the relationship between temperature and the rate of daily increase of confirmed cases of COVID-19. The multiple regression of CT^−1^ for all countries yielded a significant correlation (adjusted R^2^ = 0.610, p = 1.45 × 10^−6^) using covariates Tmin_Med, median age, land area per capita and days of cases.

To account for susceptibility variation resulting from age differences that could affect both the comorbidity levels as well as social gathering behavior, we used each country’s population median age (MA) as another covariate.(12) Because the 50 countries varied widely in size, and the density of the population could impact transmission, we used land area per capita (LAPC) as an additional covariate.(13) Finally, each country differed with respect to when it reported its identified cases, so that between the target dates of 1/22/20-4/6/20 there were different durations of time over which the course of transmission occurred. To account for this, the total number of days for each country during which they had cases of COVID-19 between 1/22/20 and 4/6/20 (Days of Cases) was used as the final covariate. In multiple regression analysis of the CT^−1^ versus the median of Tmin_Med (Table 2), we found that the rate of increase of the log of confirmed cases per unit time was significantly associated with the covariates Tmin_Med, LAPC, MA and Days of Cases (adjusted R^2^ = 0.610, p = 1.45 × 10^−6^; see Figure 2 & Table 2).

**Table 2:**
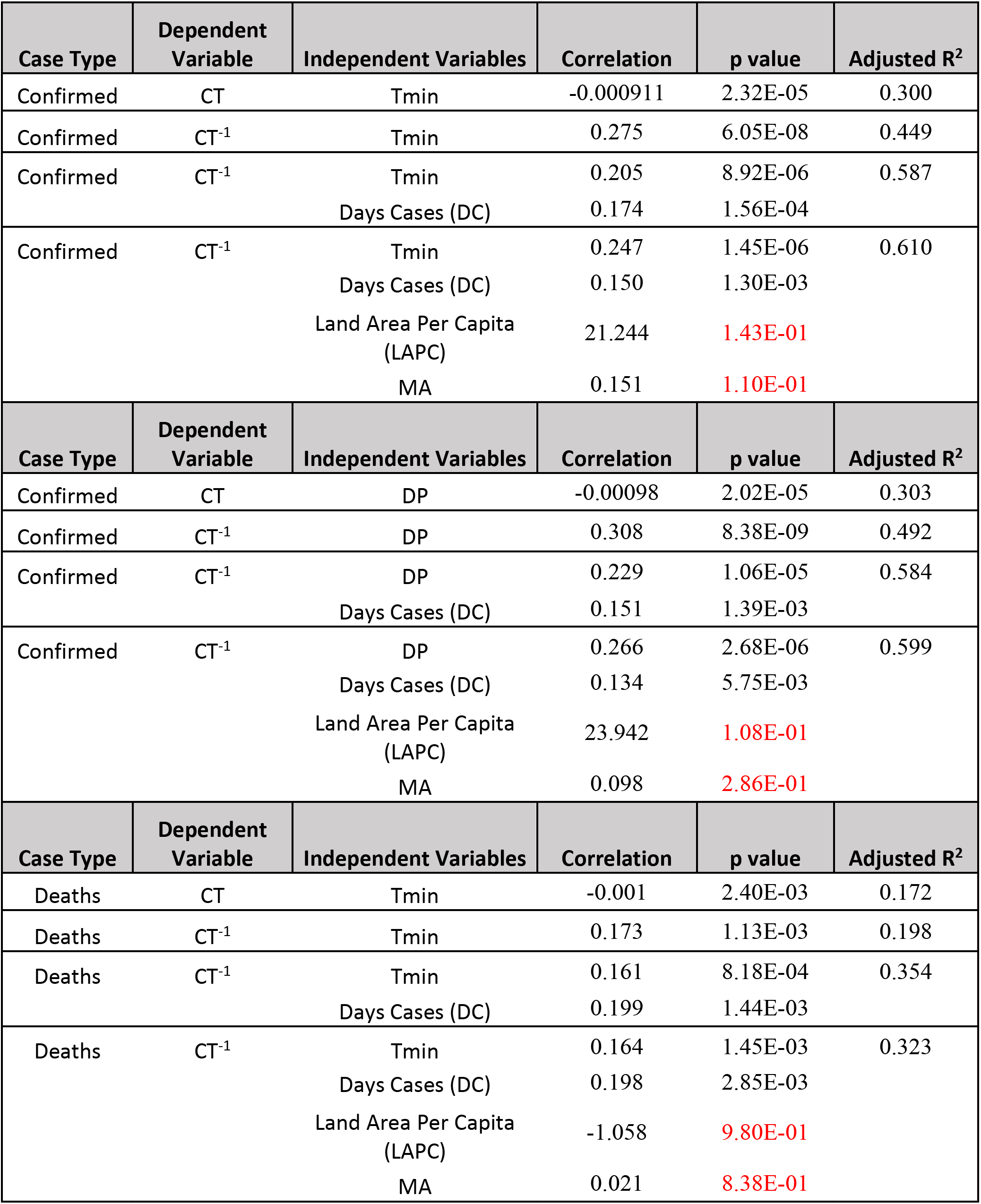
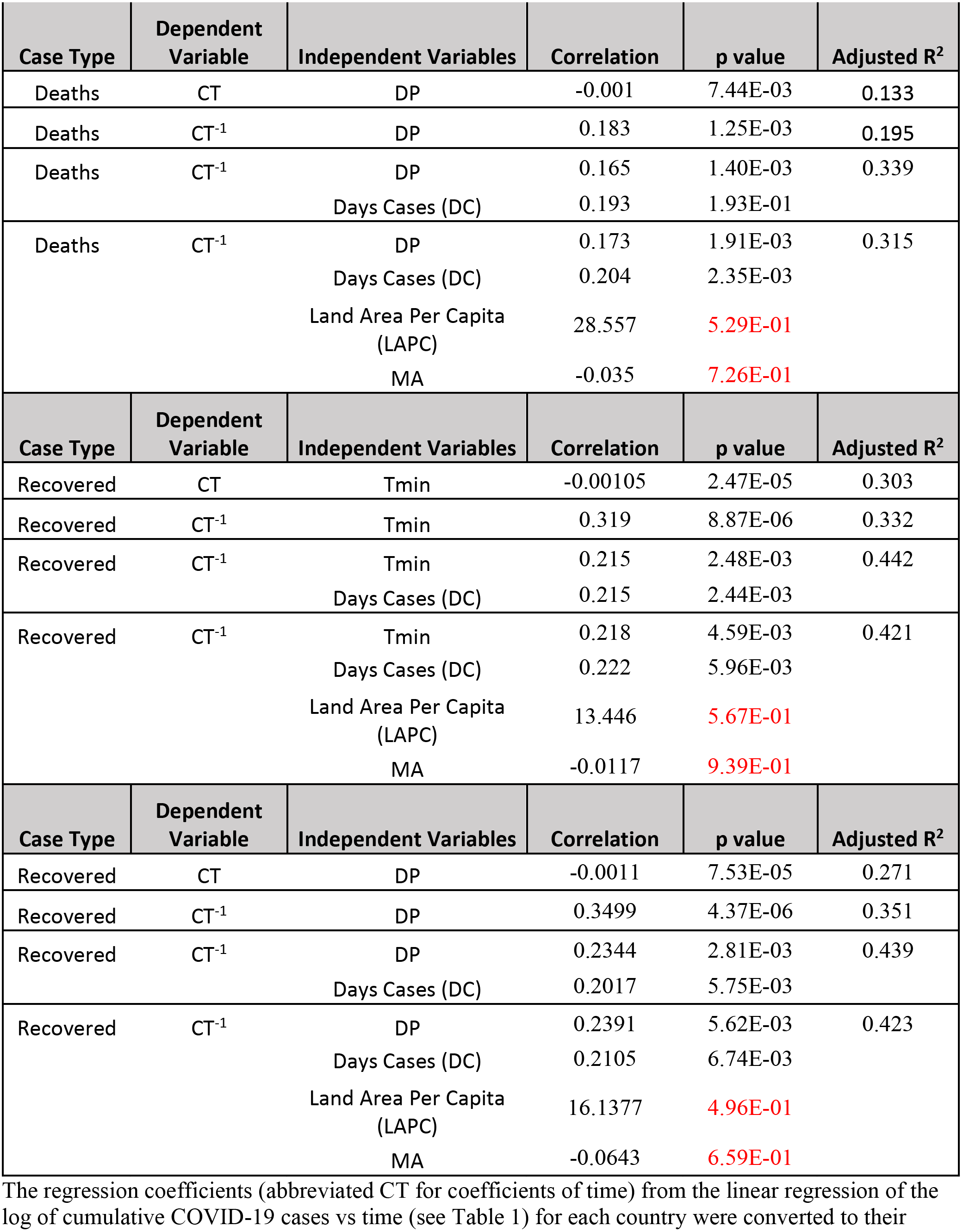
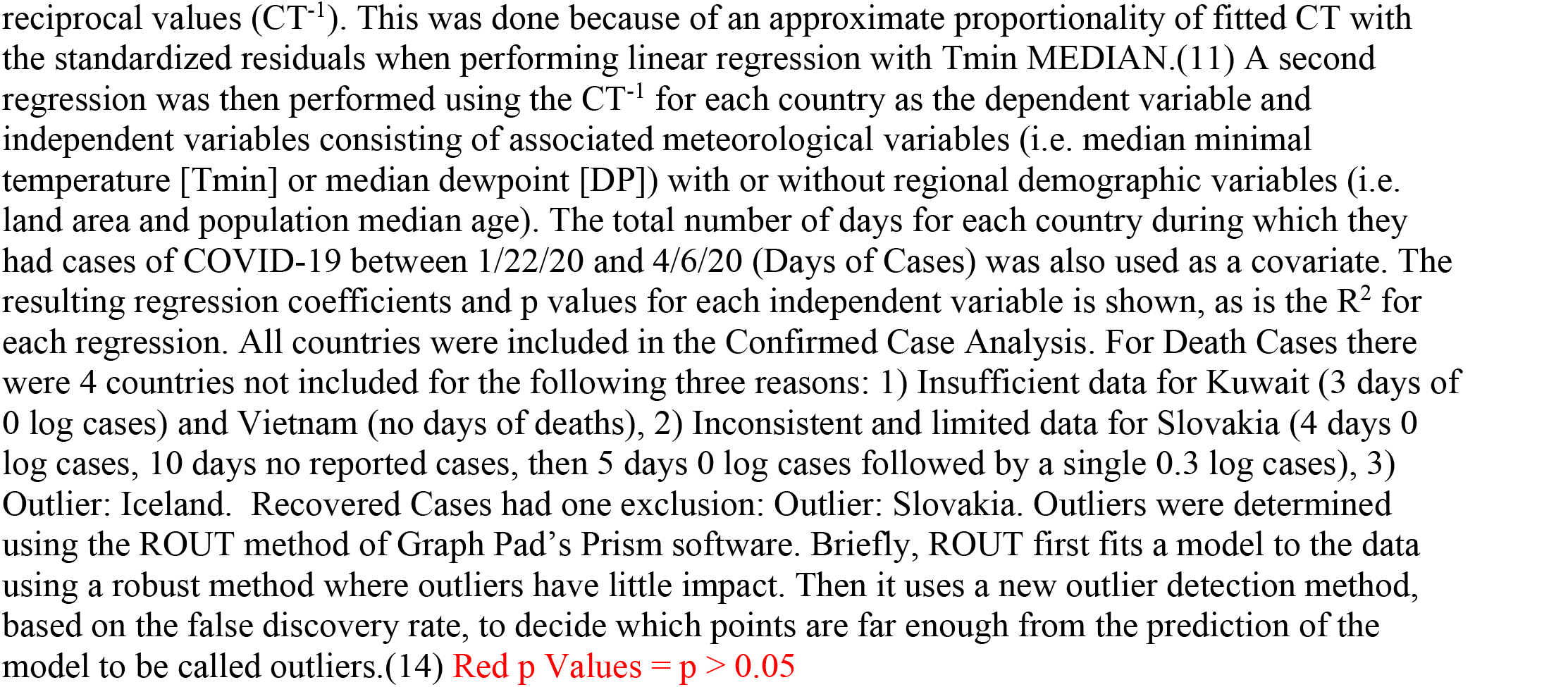
Second Regression: The association of temperature and dewpoint with the rate of rise of COVID-19 cases.

### Analysis of Dewpoint and its influence on COVID-19 rate of raise of cases

In addition to regression of CT^−1^ with Tmin_Med, we also examined the median daily DP (DP_Med) for each country. We found that the rate of increase of the log of confirmed cases per unit time using covariates DP_Med, LAPC, MA and Days of Cases showed an equally robust and statistically significant association (adjusted R^2^ = 0.599, p = 2.68 ×10^−6^). Because of the collinear relationship between Tmin_Med and DP_Med, which in linear regression resulted in an adjusted R^2^ of 0.76, (p = 1.4 × 10^−16^), we assumed that since Tmin_Med had the higher correlation with cases it was likely the primary factor underlying the association between weather and COVID-19 transmission.

### Magnitude of Effect of Temperature Change on Rate of Transmission of COVID-19

From the second regression described above, the following equation was derived:

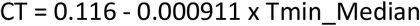

It was therefore possible to obtain CT for temperatures ranging from 30 °F to 100 °F – i.e. 0.0885 (log cases/day) to 0.0248 (log cases/day), respectively. This analysis indicated that an increase in temperature from 30 to 100 °F was associated with a 72% decrease in the rate of number of confirmed cases per day (CT 0.0885 to 0.0248), whereas a decrease in temperature from 100° to 30° F represents a 257% increase in the rate of change of confirmed cases (CT 0.0248 to 0.0885). This corresponds to 1.03% decrease in confirmed cases for each degree increase in temperature, and a 3.68% increase for each degree decrease in temperature (e.g. −72/(100-30) and 368/(100-30)).

### Correlation with rates of death and recovered cases of COVID-19

The correlation between the reciprocal of the rise in the log of the number of deaths and recovered cases of COVID-19 (CT^−1^) vs Tmin_Med was 47% and 45% less than that of confirmed cases, respectively (see Table 2). Moreover the covariates of LAPC and MA for death and recovered cases had regression p values of 0.5 or greater, suggesting that the impact of these covariates on confirmed cases was specific.

Statistical analysis was performed with StatPlus software for the Mac.(15) Graphing software employed was GraphPad Prism 8 for the Mac.(14)

## Discussion

The major finding of this study is that the rates of transmission of SARS-CoV-2 infections are strongly and robustly associated with the ambient atmospheric temperature. The results of our analysis of longitudinal data from 50 representative countries in the Northern Hemisphere suggest that the minimum daily temperature impacts both SARS CoV-2 transmissibility and viability. In addition, we found that the median daily dewpoint had a significant effect on the cumulative confirmed COVID-19 cases over time. However, because of the collinearity between dewpoint and atmospheric temperature over time, it was not possible to deduce whether the two atmospheric parameters independently affect SAR CoV-2 transmission. Since dewpoint is less predictable, and temperature more frequently monitored and used as a meteorological variable, we focused on changes in temperature for its potential predictive value.

We found that in 50 representative countries spread out over 70% of the possible latitude and longitude across the Northern Hemisphere, a slower rise in the log of daily cumulative number of confirmed cases was associated with higher daily minimum temperatures and dewpoints over the period from 1/22/20 to 4/6/20. This finding was robust, yielding an adjusted R^2^ = 0.610, p = 1.45 × 10^−6^ (using covariates Tmin_Med, median age, land area per capita, and days of cases). The correlation between the reciprocal of the rise in the log of the daily number of death and recovered cases (CT^−1^) vs Tmin_Med was almost 50% less than that of confirmed cases. This is not surprising, given that the impact of temperature on seasonal respiratory viruses has been found to have a consistent influence on the stability and transmission of infection, and not on the morbidity of infected individuals.(2) In view of these findings, atmospheric temperature would be expected to influence confirmed cases, and not deaths or the number of recovered subjects (e.g. once infected, temperature exerts a small or absent effect on the course of COVID-19 morbidity).

Although our study is a systematic and quantitative analysis of the dependence of SARS-CoV-2 infection rates on temperature and dewpoint that has demonstrated a robust and significant correlation (Table 2), it has several notable limitations. This is an ecological study and is therefore potentially subject to ecological fallacy.(16) Moreover, our data are not direct measures of individual-to-individual transmission, so cause and effect relationships cannot be established. However, the biological plausibility for our hypothesis is supported by previous work showing that the transmission rates of SARS-CoV-2 and similar viruses depend upon atmospheric temperature.(2, 17)

In temperate regions, the annual occurrence of respiratory viral diseases during the winter season--from the common cold to influenza-- has been appreciated for several thousand years.(2) A similar seasonal pattern of infections has been reported for SARS-CoV, which was prevalent mostly during winter months.(2) Despite this, there is sparse evidence on the seasonal behavior of the novel SARS-CoV-2,(3) and there are conflicting reports on how its transmission is affected by meteorological conditions. In this context, our study provides the most comprehensive and up-todate evidence for a robust and significant impact of temperature and dewpoint on SARS-CoV-2 transmissibility.

Further research is needed to clarify whether the association between temperature and SARS-CoV-2 transmissibility described here has a biological underpinning, and whether our model is accurate for longitudinal predictions of the spread of the virus over different geographical areas. Nonetheless, should the associations between temperature and the rate of transmission hold true (because of an underlying causal relationship or because of the empirical relationship described here), it may be possible to approximate the impact of weather on the rates of transmission of SARSCoV2. Because atmospheric parameters are measurable, and routinely tracked on a daily basis across the world, determining the contribution of temperature to the rate of transmission should make it possible to quantify the contribution of non-weather conditions (e.g., social and non-pharmaceutical interventions (NPI))—e.g. observed rate of transmission is equal to the contribution of meteorological variables plus the contribution of NPI. Once calibrated to the contributions of weather and social interventions and other NPI, such estimates may be able to inform and guide systematic academic, business and governmental decisions about when and to what extent to impose or relax shelter-in-place or social distancing guidelines. Such evidence-based policies may succeed in minimizing social and economic disruptions due to the pandemic while at the same time optimizing public health and safety. Finally, our analysis predicts that, between the range of 30 to 100 F, the decrease in the rate of COVID-19 transmission with increasing temperature (1% per degree F) is smaller than the increase rate of transmission with decreasing with temperatures (3.7% per degree F) in the 50 representative countries in the Northern Hemisphere countries examined. If this model is correct, then it implies that effort invested in containing, minimizing, and ideally eliminating the spread of COVID-19 during spring and summer months could pay off significantly in the fall and winter, due to potentially disproportionate effects of decreasing temperatures compared to increasing temperatures on the current pandemic.

## Data Availability

The data was all obtained from online sources referenced in the manuscript. The authors are happy to provide copies of the data that was downloaded from online sites and used in our analysis.

## Acknowledgements

The authors would like to acknowledge the intellectually stimulating discussions with Steven Babin, and the statistical consultation and mentorship provided by Howard Burkom. We also thank Jane E. Smith for her proofreading and insights.

## Bibliography

1. Astuti I, Ysrafil. Severe Acute Respiratory Syndrome Coronavirus 2 (SARS-CoV-2): An overview of viral structure and host response. Diabetes Metab Syndr. 2020;14(4):407–12.

2. Moriyama M, Hugentobler WJ, Iwasaki A. Seasonality of Respiratory Viral Infections. Annu Rev Virol. 2020.

3. Liu Y, Lam TTY, Lai FYL, Krajden M, Drews SJ, Hatchette TF, et al. Comparative seasonalities of influenza A, B and ‘common cold’ coronaviruses – setting the scene for SARS-CoV-2 infections and possible unexpected host immune interactions. J Infect. 2020.

4. Kissler SM, Tedijanto C, Goldstein E, Grad YH, Lipsitch M. Projecting the transmission dynamics of SARS-CoV-2 through the postpandemic period. Science. 2020.

5. Dong E, Du H, Gardner L. An interactive web-based dashboard to track COVID-19 in real time. The Lancet infectious diseases. 2020.

6. ISAAC STONE FISH MKS. Leaked Chinese Virus Database Covers 230 Cities, 640,000 Updates. Foreign Policy. 2020.

7. Nechepurenko I. A Coronavirus Mystery Explained: Moscow Has 1,700 Extra Deaths. New York Times. 2020.

8. Livingston E, Bucher K. Coronavirus Disease 2019 (COVID-19) in Italy. JAMA. 2020.

9. https://www7.ncdc.noaa.gov/CDO/cdoselect.cmd.

10. Eggo RM, Scott JG, Galvani AP, Meyers LA. Respiratory virus transmission dynamics determine timing of asthma exacerbation peaks: Evidence from a population-level model. Proceedings of the National Academy of Sciences. 2016;113(8):2194–9.

11. Alexopoulos EC. Introduction to multivariate regression analysis. Hippokratia. 2010;14(Suppl 1):23.

12. https://www.cia.gov/library/publications/the-world-factbook/fields/343rank.html.

13. https://www.worldometers.info/coronavirus/.

14. GraphPad. [Available from: GraphPad Prism version 8.0.0 for Mac, GraphPad Software, San Diego, California USA, www.graphpad.com.

15. StatPlus. Version v7. See https://www.analystsoft.com/en/.

16. Freedman DA. Ecological inference and the ecological fallacy. International Encyclopedia of the social & Behavioral sciences. 1999;6(4027–4030):1–7.

17. Alex W H Chin JTSC, Mahen R A Perera, Kenrie P YHui, Hui-Ling Yen, Michael C W Chan, Malik Peiris, Leo L MPoon. Stability of SARS-CoV-2 in different environmental conditions. The Lancet Microbe. 2020;Volume 1(Issue 1).

